# Willingness of urban formal sector workers to support a community-based health insurance scheme in Ethiopia

**DOI:** 10.1101/2024.11.15.24317322

**Authors:** Anagaw Derseh Mebratie, Dessalegn Shamebo, Getnet Alemu, Zemzem Shigute, Arjun S. Bedi

## Abstract

**Introduction:** The Ethiopian health system is primarily financed through household out-of-pocket expenditure and financial support from donors. High user fees lead to catastrophic health spending and limited use of services. To promote healthcare-seeking behavior and provide financial protection through enhanced domestic financing, the Ethiopian government has designed two types of health insurance schemes. These are a Community-Based Health Insurance (CBHI) scheme for the informal sector and the yet to be implemented Social Health Insurance (SHI) scheme for the formal sector. In the short run, these schemes are expected to function independently. However, in the long run it is anticipated that they will be combined, thereby pooling risk. Combining the two schemes requires solidarity across the two groups targeted by each of the schemes. Since it is likely that formal sector employees may have to bear the cost of subsidizing the CBHI scheme, this study aims to assess the extent to which formal sector employees are willing to support the CBHI scheme.

**Methods:** The paper is based on a survey of 1,919 formal sector employees and pensioners residing in the major administrative regions of the country. A survey experiment was used to elicit support for the CBHI scheme. Respondents were randomly allocated to one of five cases. These cases differed in terms of the information provided regarding the source of the CBHI subsidy and the benefits associated with the CBHI. Support for CBHI was assessed using descriptive statistics, binary and ordered logit models.

**Results:** There is strong support from urban formal sector employees for the CBHI scheme. Regardless of the scenario presented, and despite some regional variation, the key result is that at least 66% of the surveyed participants, adjusting for non-response, supported the CBHI scheme. Good knowledge of insurance increased support while existing access to health insurance lowered it.

**Conclusion:** The study provides strong evidence of solidarity and the willingness of formal sector employees to support the CBHI scheme. While this bodes well for the sustained expansion of the CBHI, it is ironic, as formal sector employees are resisting the introduction of the SHI. This reluctance stems from concerns about the costs and skepticism of the benefits of the proposed SHI, whereas the positive outcomes associated with the CBHI are widely known.

## 1. Introduction

In the past two decades, Ethiopia has made considerable progress in expanding health infrastructure and reducing child and maternal mortality rates. For instance, primary healthcare service coverage, as measured by access to a healthcare facility within a two-hour walk, increased from 51% in 2000 to greater than 90% in 2019 (FMoH, 2002; FMoH, 2021). Ethiopia is one of the few Sub-Saharan African countries which achieved the Millennium Development Goal (MDG) of reducing its under-five child mortality rate which fell from 141per 1,000 livebirths in 2000 to 50 per 1,000 live births by 2020 (World Bank, 2024). The country’s maternal mortality rate (MMR) has fallen from 9,53 in 2000 to 267 in 2020 (World Bank, 2023a). A decline which is attributed to the use of low-cost interventions such as the provision of health extension services (Rieger et al., 2019). The country has also been successful at reducing the prevalence of malaria and tuberculosis. For example, between 2000 and 2019, the number of malaria cases fell from 5.6 million to 1.8 million, and the incidence of tuberculosis fell from 400 cases to 140 cases per 100,000 people (WHO, 2023; Alene et al., 2022; WHO, 2020).

Despite such stellar progress, utilization of health care remains limited. For instance, in 2022, outpatient visits per capita was 1.09 while the WHO recommendation is between two to three out-patient visits per capita (World Bank, 2023b; FMoH, 2022a). Moreover, the country’s health system remains dependent on external donors and out-of-pocket health expenditure is substantial. In 2019-2020 about 33.5% of the health budget was covered by international donors and 30.5% of the budget was covered by individuals in the form of user fees (FMoH, 2022b). Dependency on international assistance raises the possibility that funds may not be released in a timely manner or when there are shocks that require immediate interventions (FMoH, 2010; ZMOH, 2017; Nyamugira et al., 2022). At the same time, reliance on out-of-pocket health payments leads to catastrophic health expenditure which has impoverishing effects such as depletion of productive assets, reduction in consumption, forgone human capital investment (O’Donnell et al., 2005; De Weerdt and Dercon, 2006; Flores et al., 2008; Karami et al., 2009).

Since 2008, to address these issues and specifically to mobilize domestic resources to finance healthcare and expand access to quality care, the Ethiopian government introduced several healthcare financing reforms (Abt Associates, 2012). One of the components of these reforms was the introduction of pre-payment insurance schemes. Specifically, a voluntary CBHI scheme for the informal sector and a still to be implemented, mandatory SHI scheme for the formal sector.^1^ In 2011, a pilot CBHI scheme was rolled out in 13 selected districts, and by 2023, the scheme had expanded to 1,011 districts and towns covering about 87.4% of the districts and town administrations in the country and about 12.1 million households.^2^ According to the Ethiopian Health Insurance Service (EHIS), in 2023, scheme enrollment rate was 78% and renewal was 91%.

As compared to other Sub-Saharan African countries, the scheme has been successful (Mebratie et al., 2015; Alemu et al. 2024).^3^ Impact evaluation studies show that the scheme has enhanced access to healthcare services and reduced the incidence of borrowing (Mebratie et al., 2013; Yilma et al., 2015). While the scheme does not provide complete financial protection as households still have to pay for healthcare services due to limited knowledge of scheme design or lack of drug or laboratory equipment at health facilities, it has certainly alleviated the financial burden (Mebratie et al., 2015).

While the CBHI scheme has been scaled up, its sustainability requires strong and continued political and financial support from the government. Consistent with the basic insurance principle of risk pooling whereby larger risk pools enhance scheme sustainability (Zweifel et al., 2009), the long-term aim of the government is to combine the CBHI and the still to be introduced SHI. This arrangement is expected to spread risk, enable cross-subsidization of financial resources from the formal to the informal sector and ensure sustained access to health care regardless of socio-economic status (Douwes et al., 2018; Harris et al., 2011; Buchmueller & Couffinhal, 2004). Given the expected financial burden on the formal sector, a prerequisite for combining the two health care insurance schemes is solidarity across the two groups targeted by the schemes. Accordingly, this study aims to assess the extent to which formal sector employees are willing to support the CBHI.

To realize it’s objectives, a survey of 1,919 formal sector employees and pensioners residing in the major administrative regions of the country was conducted. A survey experiment was used to elicit support for the CBHI scheme. Respondents were randomly allocated to one of five cases. These cases differed in terms of the information provided on the source of the CBHI subsidy and the benefits associated with it. After receiving information, respondents were asked to signal their level of support for the scheme.

To preview our results, we find that there is strong support from urban formal sector employees for the CBHI scheme. Regardless of the scenario presented, and despite some regional variation, the key result is that about 76% of the surveyed participants support the CBHI scheme. Such a high level of support suggests that, when the time comes, it should be possible to combine the two schemes without substantial friction. Such evidence is clearly useful in informing the efforts of the government and the EHIS as they strive to meet the country’s goal of developing a sustainable health insurance system.

## 2. The design of health insurance schemes in Ethiopia – a brief description

In 2008, in collaboration with development partners, the Ethiopian Federal Ministry of Health prepared a health insurance strategy to establish two types of health insurance schemes. These were a CBHI scheme to cater for rural and informal sector workers in urban centers and a SHI scheme for formal sector employees and pensioners. International experience and local feasibility studies informed scheme design.^4^ Subsequently, the CBHI was piloted in 13 districts in 2011 and the scheme was evaluated in 2013 (HFG, 2015). Based on the evaluation, the initial design of the scheme was revised and it was scaled up in 2016.^5^ The basic design features of the Ethiopian CBHI and SHI are described below.

### 2.1 Community based health insurance scheme

The CBHI is a voluntary health insurance scheme offered to rural workers and informal sector workers in urban areas.^6^ While district and regional governments are responsible for the implementation and administration of the CBHI schemes, community members are involved in management and evaluation. Specific tasks for community members include community mobilization, premium collection, membership renewal and scheme monitoring.

Enrolment in the scheme is only possible at the household level and includes coverage for core family members (wife, husband, and children under 18).^7^ Other household members may be covered based on additional payment. At inception, the premium amount and the frequency of payment was sensitive to local contexts and varied across regions.^8^ However, since 2017, greater uniformity has been imposed and the premium also varies depending on household size. In rural areas, depending on household size, the annual premium amounts to between ETB 800 and ETB 1,100 per annum and in urban areas between ETB 900 and ETB 3500 (ANRS, 2024).^9^ While the monthly contribution may be considered modest as compared to the expected benefits, the scheme still includes a fee waiver system for indigent groups - up to 10% of the eligible households in a district may enroll in the scheme as fee waiver beneficiaries.^10^ To enhance financial sustainability and retain affordability, the federal government provides a general subsidy to support the CBHI. Since 2015 this subsidy was reduced from 25% to 10% of the total premiums collected by the CBHI scheme (Abt Associates, 2015; Segahu, 2018; EHIA, 2020). While the bulk of the schemes pool risk at the district level, in some parts of the country risk is pooled at a higher level of aggregation.^11^

Enrolled members and their families are entitled to inpatient and outpatient care from public sector health facilities (FDRE, 2012; SIAPS, 2016). There are no deductibles and co-payments and payment is not required at point-of-use. Care at private health facilities and medical treatment abroad is not covered. The scheme follows a gatekeeper system and scheme members first need to visit the nearest health center and subsequently they may be referred to tertiary care facilities. The benefit package excludes health services such as those related to drug abuse, traffic accidents, occupational injuries, cosmetic surgeries, organ transplants, eyeglasses, and contact lenses. The scheme does not provide financial coverage for treatment of injuries resulting from social unrest and natural disasters (FDRE, 2012; SIAPS, 2016).

### 2.2 Social Health Insurance Scheme

The legal and administrative aspects of the SHI scheme have been established, and the Ethiopian Health Insurance Service (EHIS) has been created to manage the scheme (Council of Ministers Regulation No. 271/2012). Operational documents have been prepared, and in 2014, the EHIS – agency established to manage the schemes – setup branch offices in selected towns (FDRE, 2012, Abt Associates, 2015; SIAPS, 2016).

Despite all the background preparations, the scheme has not yet been implemented. While the actual policy may differ from the initial design, at the moment, the proposed SHI is expected to be introduced in one go and enrolment is mandatory for all formal sector (private, public, NGO, pensioners) employees. The SHI proclamation sets the premium at 6% of an employees’ gross salary and requires an equal contribution (3% each) from both the employee and the employer. For pensioners, the premium is set at 2% of their pension amount with 1% to be covered by the pensioner and a government contribution of 1% (FDRE, 2012; Ali, 2014). The benefit package is the same as in the case of the CBHI, that is, enrolled members and their families are entitled to inpatient and outpatient care from public sector health facilities (FDRE, 2012; SIAPS, 2016).

While scheme launch has often seemed imminent, it has been delayed, mainly for two reasons. First, formal sector employees have expressed their unwillingness to pay the proposed SHI premium. Although there are no national-level studies, formal willingness to pay (WTP) studies conducted on various samples of civil servants in Addis Ababa show that between 17% to 35% of the sampled respondents are willing to pay the premium with a mean WTP ranging between 1.5% to 2.5% of gross monthly salaries (Zarepour et al. 2023). A meta-analysis covering 18 studies showed that, on average, about 42% of formal sector workers are WTP for SHI (Bayked et al. 2023). The second reason for the delay is the scheme’s healthcare coverage. The scheme is likely to restrict coverage to public health facilities rather than covering potentially higher quality private care. This restriction is likely to drive up costs of health care as it requires SHI enrollees to pay the premium and yet they may continue to seek care at private health facilities.^12^ A recent paper (Zarepour et al. 2023) on healthcare seeking behavior of formal sector workers shows that restricting access to public health facilities is at odds with the healthcare seeking behavior of formal sector employees, especially for outpatient care.

## 3. Data and Methods

### 3.1 Data

This paper is based on a retrospective cross-sectional survey conducted in-person between June and July 2016. The survey was canvassed in four of the country’s main cities. These included Addis Ababa – the country’s capital, Bahir Dar, the largest city in the Amhara region, Hawassa, the largest city in the SNNP region, and Mekelle, the largest city in the Tigray region. These cities were purposively selected as they accounted for 20% of the estimated 4.3 million formal sector employes in the country. To obtain a representative sample and include those targeted by the SHI, the survey covered five categories of employees - civil servants, public sector enterprise employees, private sector workers, NGO workers, pensioners (former civil servant and public sector enterprise workers) and the distribution of the sample across cities and type of formal sector employees was determined based on the distribution of the population of formal sector workers in these cities. Population level information was obtained from the Ethiopian Ministry of Civil Service, the Central Statistical Agency, the Ethiopian Chamber of Commerce and Sectorial Associations, and the Ethiopian social security agency. Considering the budget, a sample size of 2,100 respondents was targeted (for additional details on the sampling, see Zarepour et al. 2023).

After ensuring that a selected respondent was willing to participate in the survey, enumerators gathered individual and household-level information. The survey contained a household roster which gathered socio-economic information on all household members, their health status and lifestyle choices, outpatient and inpatient health care utilization, financing of heath care, understanding of health insurance and whether they currently had any form of health insurance. Specifically, for the purposes of the current paper, a survey-based experiment was used to gather information on respondent support or lack thereof for the CBHI. Details are provided in the subsequent section.

In total, while a little less than the targeted figure, the survey was able to cover 1,919 respondents distributed across the four cities and five types of employees (see Table A1). As shown in Table A1, the distribution of the sample across types of employees is very similar to their distribution in the population. While not perfect, arguably the sample is representative of formal sector employees in urban Ethiopia.

### 3.2 Support for the CBHI amongst formal sector employees – a survey-based experiment

To elicit support for the CBHI amongst formal sector employees, the paper relies on a survey-based experiment. Survey respondents were *randomly* assigned to one of five different cases. Each case had two questions (see Table 1, and Table A2). The first question in each case was whether respondents had heard of the CBHI scheme. The second question, which was posed after providing information on different ways of subsidizing the CBHI and/or providing information on benefits accruing due to the CBHI, asked respondents whether they opposed or supported the scheme using a five-point Likert scale. To guard against “cheap talk”, those who stated that they (strongly) supported the CBHI scheme were asked to sign a slip of paper to confirm their level of commitment endorsing continued government support of the CBHI scheme.

**Table 1.** Alternative scenarios presented to survey respondents.

|  | **Source of subsidy** | **CBHI benefits/effectiveness** | *N* |
| --- | --- | --- | --- |
| Case 1 | No information | No information | 386 |
| Case 2 | Formal sector workers | No information | 344 |
| Case 3 | International donors | No information | 340 |
| Case 4 | Formal sector workers | The scheme is effective in terms of coverage, creating access to care and providing financial protection | 325 |
| Case 5 | No information | The scheme is effective in terms of coverage, creating access to care and providing financial protection | 267 |

**Notes:** Of the 1,919 respondents, 257 (13%) did not provide responses to the question on support for the CBHI. Table A2 provides details on the information shared in the cases.

As displayed in Table 1 and in more detail in Table A2, the first case did not provide any information but simply asked respondents whether they support the CBHI scheme (baseline case). The second case explicitly stated that the cost of subsidizing the scheme would be borne by those working in the formal sector. The third case stated that the subsidy would be covered by international donors. The fourth case indicated that the subsidy would be covered by taxpayers but provided research-based information on scheme effectiveness in terms of enhancing access to health care and reducing OOP health expenditure. The final case did not contain any information on how the subsidy would be financed but provided the same information on scheme effectiveness as in case four.

As compared to the baseline case, the expectation is that support for the CBHI scheme or in other words, solidarity with rural and informal sector workers, will be lower when it is explicitly pointed out that formal sector workers subsidize the CBHI (Case 2) and higher as compared to the baseline case when the subsidy is financed by external sources (Case 3). The lower support for the CBHI scheme (Case 2) is expected to be ameliorated when research-based information is provided on the effectiveness of the scheme (Case 4). Support for CBHI in Case 5 is expected to be higher than the baseline. The rationale for the variations across cases is based on the main concerns highlighted in Section 2, that is, maintaining and enhancing the affordability of the CBHI requires continued government financial support and at the same time there is skepticism on the quality and usefulness of the health services which may be availed through the CBHI and eventually through the SHI.

As mentioned above, each of the respondents was randomly assigned to *only one* of the cases. Due to this randomization, there is no reason to expect that the sociodemographic and other traits of respondents differ across the five cases. Therefore, it should be possible to identify the effect of the variations in the cases on the level of support for the CBHI. In other words, while the sociodemographic traits of respondents may influence their support for the CBHI, their traits are not likely to influence the effect of the cases on support for the CBHI. Formal statistical tests clearly show that the profiles of the study participants do not differ across the five cases (Table A3).

### 3.3 Methods

We use descriptive statistics to provide information on the sample respondents and to examine their understanding of health insurance. Subsequently, we probe whether respondents have heard of the CBHI and their level of support for the scheme. Finally, we examine willingness to support the scheme as a function of the characteristics of the case offered. To do so we estimate binary logit models (support, do not support) and an ordered logit model where the outcome variable has four options (strongly opposed or opposed, neither support nor opposed, support, and strongly support).^13^ We estimate versions of the logit models which control only for the cases proposed to the respondents and a specification which also controls for respondents’ sociodemographic characteristics (sex, age, sector of employment, educational and marital status, religion and ethnicity, income, house ownership, family size), health related traits (household health care use, life style of family members – incidence of drinking, smoking, chewing chat, health insurance, knowledge of health insurance) and a set of geographical indicator variables.

## 4. Results

### 4.1 Descriptive statistics

The sample consists of 1,919 individuals and as shown in Table A1, the distribution of the sample across sectors matches their distribution in the population. The sample consists of 33% public sector workers, about 10% work in public enterprises, 41.4% are engaged in the private sector/NGO while the remainder (15%) are pensioners. About 64% of the sample is male. Since the sample consists of formal sector workers it is not surprising that about 72% have tertiary education. Amongst those still working, education levels are highest amongst public sector workers – 87% have tertiary education as opposed to about 70% amongst private/NGO sector workers. Regarding monthly income, pensioners have the lowest income while workers in public sector enterprises record the highest income (Table 2).

**Table 2.**
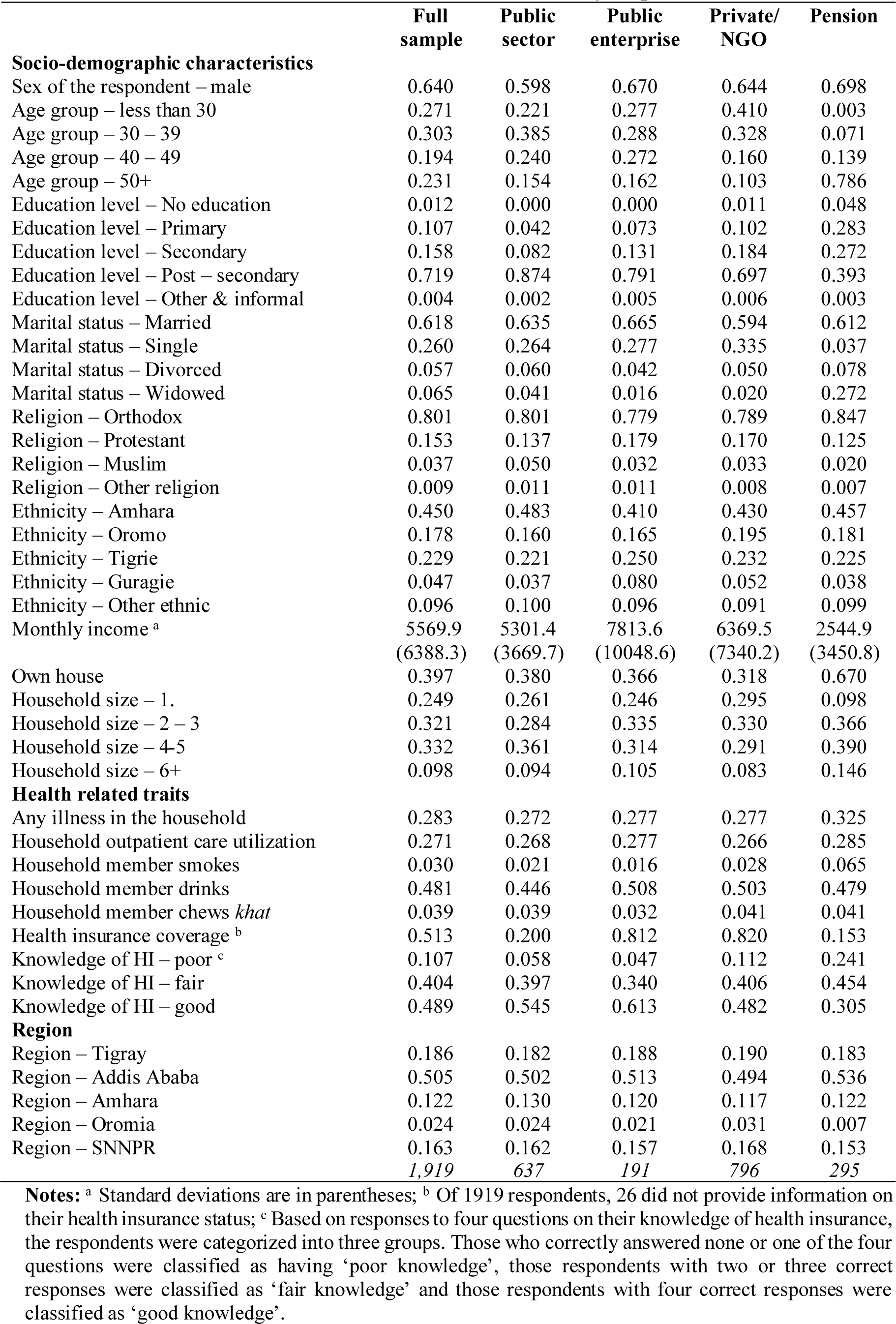
Socioeconomic characteristics of survey respondents.

|  | Full sample | Public sector | Public enterprise | Private/ NGO | Pension |
| --- | --- | --- | --- | --- | --- |
| **Socio-demographic characteristics** |  |  |  |  |  |
| Sex of the respondent – male | 0.640 | 0.598 | 0.670 | 0.644 | 0.698 |
| Age group – less than 30 | 0.271 | 0.221 | 0.277 | 0.410 | 0.003 |
| Age group – 30 – 39 | 0.303 | 0.385 | 0.288 | 0.328 | 0.071 |
| Age group – 40 – 49 | 0.194 | 0.240 | 0.272 | 0.160 | 0.139 |
| Age group – 50+ | 0.231 | 0.154 | 0.162 | 0.103 | 0.786 |
| Education level – No education | 0.012 | 0.000 | 0.000 | 0.011 | 0.048 |
| Education level – Primary | 0.107 | 0.042 | 0.073 | 0.102 | 0.283 |
| Education level – Secondary | 0.158 | 0.082 | 0.131 | 0.184 | 0.272 |
| Education level – Post – secondary | 0.719 | 0.874 | 0.791 | 0.697 | 0.393 |
| Education level – Other & informal | 0.004 | 0.002 | 0.005 | 0.006 | 0.003 |
| Marital status – Married | 0.618 | 0.635 | 0.665 | 0.594 | 0.612 |
| Marital status – Single | 0.260 | 0.264 | 0.277 | 0.335 | 0.037 |
| Marital status – Divorced | 0.057 | 0.060 | 0.042 | 0.050 | 0.078 |
| Marital status – Widowed | 0.065 | 0.041 | 0.016 | 0.020 | 0.272 |
| Religion – Orthodox | 0.801 | 0.801 | 0.779 | 0.789 | 0.847 |
| Religion – Protestant | 0.153 | 0.137 | 0.179 | 0.170 | 0.125 |
| Religion – Muslim | 0.037 | 0.050 | 0.032 | 0.033 | 0.020 |
| Religion – Other religion | 0.009 | 0.011 | 0.011 | 0.008 | 0.007 |
| Ethnicity – Amhara | 0.450 | 0.483 | 0.410 | 0.430 | 0.457 |
| Ethnicity – Oromo | 0.178 | 0.160 | 0.165 | 0.195 | 0.181 |
| Ethnicity – Tigrie | 0.229 | 0.221 | 0.250 | 0.232 | 0.225 |
| Ethnicity – Guragie | 0.047 | 0.037 | 0.080 | 0.052 | 0.038 |
| Ethnicity – Other ethnic | 0.096 | 0.100 | 0.096 | 0.091 | 0.099 |
| Monthly income <sup>a</sup> | 5569.9<br>(6388.3) | 5301.4<br>(3669.7) | 7813.6<br>(10048.6) | 6369.5<br>(7340.2) | 2544.9<br>(3450.8) |
| Own house | 0.397 | 0.380 | 0.366 | 0.318 | 0.670 |
| Household size – 1. | 0.249 | 0.261 | 0.246 | 0.295 | 0.098 |
| Household size – 2 – 3 | 0.321 | 0.284 | 0.335 | 0.330 | 0.366 |
| Household size – 4-5 | 0.332 | 0.361 | 0.314 | 0.291 | 0.390 |
| Household size – 6+ | 0.098 | 0.094 | 0.105 | 0.083 | 0.146 |
| **Health related traits** |  |  |  |  |  |
| Any illness in the household | 0.283 | 0.272 | 0.277 | 0.277 | 0.325 |
| Household outpatient care utilization | 0.271 | 0.268 | 0.277 | 0.266 | 0.285 |
| Household member smokes | 0.030 | 0.021 | 0.016 | 0.028 | 0.065 |
| Household member drinks | 0.481 | 0.446 | 0.508 | 0.503 | 0.479 |
| Household member chews *khat* | 0.039 | 0.039 | 0.032 | 0.041 | 0.041 |
| Health insurance coverage <sup>b</sup> | 0.513 | 0.200 | 0.812 | 0.820 | 0.153 |
| Knowledge of HI – poor <sup>c</sup> | 0.107 | 0.058 | 0.047 | 0.112 | 0.241 |
| Knowledge of HI – fair | 0.404 | 0.397 | 0.340 | 0.406 | 0.454 |
| Knowledge of HI – good | 0.489 | 0.545 | 0.613 | 0.482 | 0.305 |
| **Region** |  |  |  |  |  |
| Region – Tigray | 0.186 | 0.182 | 0.188 | 0.190 | 0.183 |
| Region – Addis Ababa | 0.505 | 0.502 | 0.513 | 0.494 | 0.536 |
| Region – Amhara | 0.122 | 0.130 | 0.120 | 0.117 | 0.122 |
| Region – Oromia | 0.024 | 0.024 | 0.021 | 0.031 | 0.007 |
| Region – SNNPR | 0.163 | 0.162 | 0.157 | 0.168 | 0.153 |
|  | 1,919 | 637 | 191 | 796 | 295 |

**Notes:** <sup>a</sup> Standard deviations are in parentheses; <sup>b</sup> Of 1919 respondents, 26 did not provide information on their health insurance status; <sup>c</sup> Based on responses to four questions on their knowledge of health insurance, the respondents were categorized into three groups. Those who correctly answered none or one of the four questions were classified as having ‘poor knowledge’, those respondents with two or three correct responses were classified as ‘fair knowledge’ and those respondents with four correct responses were classified as ‘good knowledge’.

Table 2 also provides information on the current health insurance status and knowledge of health insurance of sample respondents. On average, about 51% of respondents have health insurance. This figure varies substantially across sectors with 81-82% of public sector enterprise and private sector/NGO workers reporting that they have health insurance while the corresponding figures are 20% among public sector workers and 15% amongst pensioners. These sharp difference in coverage supports the idea that enterprises, both public and private, and NGOs offer health insurance to attract workers while those in the public sector are not offered similar benefits (Obse et al., 2016; FMoH, 2022a). While almost all respondents had heard of health insurance (92%), a much smaller fraction – about 49% - were able to provide correct answers to a set of four questions on the functioning of insurance (see Tables 2 and 3). There is some variation across sectors with “good knowledge” of insurance varying from a low of 30.5 percent amongst pensioners to a high of 61.3 percent amongst private sector/NGO workers.

**Table 3.** Understanding of health insurance (%)

|  | Correct | Incorrect | Do not know | N |
| --- | --- | --- | --- | --- |
| Only those who fall sick should consider paying for (HI)? | 2.24 | 92.43 | 5.32 | 1,916 |
| HI programs are like savings scheme, you will receive interest and get your money back? | 14.81 | 64.68 | 20.50 | 1,917 |
| In HI programs you pay money (premiums) for the HI to finance your future health care needs? | 81.32 | 7.67 | 11.01 | 1,917 |
| If you do not claim any costs through HI your premium will be returned? | 11.22 | 68.44 | 20.34 | 1,917 |

### 4.2 Awareness and willingness to support CBHI

Of the 1,919 respondents, about 13% (257) refused to respond to the cases. The non-respondents tend to be older, less educated, more likely to be in the lower income categories, and more likely to reside in Tigray (see Table A4). The reasons for their lack of response is unclear but it could indicate lack of support for the CBHI, thereby providing an inflated idea of support for the scheme. We return to this issue in the discussion. Conditional on (strongly) supporting the scheme, almost all (97%) signed a piece of paper indicating their commitment.

In response to the first question on scheme awareness, regardless of the case, almost all (97%) the respondents had heard about the scheme (Table 4). Subsequently, all those who had heard about the CBHI were asked to indicate their support or lack thereof, for it. As shown in Table 4, regardless of whether the case provided no information, or provided information on the source of subsidy (domestic or international support) or information on the subsidy and research-based findings on the benefits that the CBHI has yielded or only information on the benefits, there was strong overall support for the CBHI. Regardless of the case, about 76% of the respondents (strongly) supported the CBHI while 11% were indifferent and 13% opposed the scheme. While there are no statistically significant differences in support across the cases, there are two notable points. First, regardless of whether formal sector workers are expected to subsidize the scheme or not they do not waver in terms of their support (compare cases 1 and 2). Second, there is greater support when research findings on benefits are provided - 73% versus 79% (compare cases 1 and 5).

**Table 4.** Awareness and Support for the CBHI Scheme.

|  | Case 1 | Case 2 | Case 3 | Case 4 | Case 5 | Kruskal–Wallis test (P-value) | Total |
| --- | --- | --- | --- | --- | --- | --- | --- |
| Heard about the CBHI scheme (%) | 96.6 | 96.8 | 97.9 | 98.2 | 97.0 | 0.6456 | 97.3 |
| N | (373) | (333) | (333) | (319) | (259) |  | (1617) |
| Opposed | 13.0 | 14.2 | 13.5 | 13.9 | 12.0 | 0.7203 | 13.4 |
| N | (50) | (49) | (46) | (45) | (32) |  | (222) |
| Neither opposed nor support | 14.3 | 8.1 | 10 | 12.6 | 9.0 |  | 11.0 |
| N | (55) | (28) | (34) | (41) | (24) |  | (182) |
| Support | 52.3 | 57.0 | 59.1 | 54.5 | 58.1 |  | 56.0 |
| N | (202) | (196) | (201) | (177) | (155) |  | (931) |
| Strongly support | 20.5 | 20.6 | 17.4 | 19.1 | 21.0 |  | 19.7 |
| N | (79) | (71) | (59) | (62) | (56) |  | (327) |

To investigate the traits of those who support/do not support the scheme, Table 5 provides marginal effects from a logit model. The table presents four sets of estimates. Consistent with the patterns noted in Table 4, the level of support does not vary between the baseline and cases two, three and four. However, there is a statistically significant 6 percentage point higher support amongst respondents who are provided with research-based findings on the benefits associated with CBHI (Table 5, model 1). The inclusion of several traits reduces the statistical significance of the cases variables but the qualitative story remains the same. The additional information yielded by these specifications is that individuals who already have insurance are less likely to support CBHI (6 percentage point effect). Those respondents who have greater knowledge of insurance are more likely to support the CBHI – the effect is 12-16 percentage points higher depending on the model and whether an individual has fair or good knowledge of health insurance. Finally, there is large regional variation with the greatest support for the CBHI amongst formal sector workers in SNNPR, followed by Amhara and Addis Ababa (model 3). The lowest level of support in Tigray followed by the Oromia region.

**Table 5.** Probability of supporting CBHI – marginal effects from binary logit models.

| VARIABLES<br>/ | Model 1 | Model 2 | Model 3 | Model 4 |
| --- | --- | --- | --- | --- |
| **Scenario** |  |  |  |  |
| Case two – formal sector subsidizes,<br>no information on scheme benefits (ref: Case one) | 0.0457<br>(0.0292) | 0.0434<br>(0.0293) | 0.0456<br>(0.0287) | 0.0487*<br>(0.0287) |
| Case three – international donor subsidizes,<br>no information on scheme benefits | 0.0346<br>(0.0296) | 0.0336<br>(0.0297) | 0.0296<br>(0.0295) | 0.0314<br>(0.0297) |
| Case four – formal sector subsidizes,<br>information on scheme benefits provided | 0.00688<br>(0.0308) | 0.00483<br>(0.0310) | 0.000846<br>(0.0309) | 0.00230<br>(0.0310) |
| Case five – no information on subsidizer,<br>information on scheme benefits provided | 0.0590*<br>(0.0304) | 0.0578*<br>(0.0305) | 0.0417<br>(0.0313) | 0.0445<br>(0.0313) |
| **Insurance membership and knowledge** |  |  |  |  |
| Insurance membership (ref: non-members) | . | . | . | -0.0604**<br>(0.0298) |
| Knowledge of health insurance is fair<br>(ref: poor knowledge) | . | 0.128***<br>(0.0341) | 0.125***<br>(0.0340) | 0.134***<br>(0.0343) |
| Knowledge of health insurance is good | . | 0.117***<br>(0.0364) | 0.146***<br>(0.0367) | 0.155***<br>(0.0371) |
| **Sector** |  |  |  |  |
| Sector-Public enterprises (ref: public service) | . | 0.0167<br>(0.0362) | 0.0140<br>(0.0359) | 0.0515<br>(0.0363) |
| Sector-Private and NGO | . | 0.00445<br>(0.0250) | 0.00382<br>(0.0247) | 0.0452<br>(0.0308) |
| Sector-Pension | . | 0.0575<br>(0.0363) | 0.0545<br>(0.0356) | 0.0556<br>(0.0356) |
| **Region** |  |  |  |  |
| Region – SNNPR (ref: Tigray) | . | . | 0.201***<br>(0.0219) | 0.197***<br>(0.0228) |
| Region – Addis Ababa | . | . | 0.0939***<br>(0.0291) | 0.103***<br>(0.0295) |
| Region – Amhara | . | . | 0.110***<br>(0.0286) | 0.0985***<br>(0.0310) |
| Region – Oromia | . | . | 0.0657<br>(0.0551) | 0.0772<br>(0.0526) |
| *Observations* | *1,662* | *1,662* | *1,662* | *1,646* |

**Notes:** \*\*\* p<0.01, \*\* p<0.05, \* p<0.1. Standard errors are in parentheses. We explored specifications with all the individual and household controls listed in Table 2. In the interests of brevity, we report coefficients on a selected set of relevant variables. The variables that are not displayed were statistically insignificant.

As a final step, instead of a two-part (support/do not support specification) we estimated an ordered logit with four categories - ranging from opposing the CBHI to strong support (see Table 6). These estimates yield subtle insights. Like the results in Tables 4 and 5, there is no indication that the type of information provided about the design features of the CBHI has a bearing on the support expressed by formal sector employees. As in the case of the binary logit model, knowledge of health insurance has a bearing on support for the CBHI scheme. Employees with good knowledge of health insurance are 10 to 11 percentage points more likely to strongly support the development of the CBHI scheme compared to those with poor knowledge. Those with health insurance are not just neutral but more likely to oppose support for the CBHI. Similarly, while regional disparities remain salient, formal sector workers in Tigray and Oromia actively oppose support for the CBHI as opposed to being neutral.

**Table 6.** Factors affecting support for CBHI scheme, marginal effects after ordered logit.

| VARIABLES | Model 1 |  |  |  | Model 2 |  |  |  |
| --- | --- | --- | --- | --- | --- | --- | --- | --- |
|  | Oppose | Neither oppose<br>nor support | Support | Strongly support | Oppose | Neither oppose<br>nor support | Support | Strongly<br>support |
| **Scenario** |  |  |  |  |  |  |  |  |
| Case two (ref. case one) | -0.0131<br>(0.0166) | -0.00774<br>(0.0098) | 0.00294<br>(0.0039) | 0.0179<br>(0.0226) | -0.0121<br>(0.0165) | -0.00690<br>(0.00945) | 0.00301<br>(0.00422) | 0.0160<br>(0.0219) |
| Case three | 3.05E-5<br>(0.0165) | 1.80E-05<br>(0.0097) | -6.85E-6<br>(0.0037) | -4.17E-05<br>(0.0225) | 0.00401<br>(0.0165) | 0.00229<br>(0.00941) | -0.00100<br>(0.00412) | -0.00530<br>(0.0218) |
| Case four | 0.00327<br>(0.0167) | 0.00193<br>(0.0099) | -0.00073<br>(0.0038) | -0.00447<br>(0.0229) | 0.00940<br>(0.0167) | 0.00536<br>(0.00953) | -0.00234<br>(0.00420) | -0.0124<br>(0.0221) |
| Case five | -0.0204<br>(0.0177) | -0.0121<br>(0.0105) | 0.00458<br>(0.0043) | 0.0279<br>(0.0242) | -0.0136<br>(0.0177) | -0.00774<br>(0.0101) | 0.00338<br>(0.00454) | 0.0179<br>(0.0234) |
| **Insurance membership and knowledge** |  |  |  |  |  |  |  |  |
| Insurance membership (ref: non-members) | . | . | . | . | 0.0258*<br>(0.0156) | 0.0147*<br>(0.00887) | -0.00643<br>(0.00431) | -0.0341*<br>(0.0205) |
| Knowledge on health insurance is fair<br>(ref: poor knowledge) | . | . | . | . | -0.0772***<br>(0.0204) | -0.0441***<br>(0.0117) | 0.0192***<br>(0.00732) | 0.102***<br>(0.0269) |
| Knowledge on health insurance is good | . | . | . | . | -0.0866***<br>(0.0207) | -0.0494***<br>(0.0118) | 0.0216***<br>(0.00786) | 0.114***<br>(0.0271) |
| **Sector** |  |  |  |  |  |  |  |  |
| Sector-Public enterprises (ref: public service) | . | . | . | . | -0.0104<br>(0.0217) | -0.00591<br>(0.0124) | 0.00258<br>(0.00545) | 0.0137<br>(0.0286) |
| Sector-Private and NGO | . | . | . | . | -0.0185<br>(0.0165) | -0.0106<br>(0.00943) | 0.00461<br>(0.00436) | 0.0245<br>(0.0218) |
| Sector-Pension | . | . | . | . | -0.0233<br>(0.0213) | -0.0133<br>(0.0122) | 0.00580<br>(0.00557) | 0.0308<br>(0.0281) |
| **Region** |  |  |  |  |  |  |  |  |
| Region – SNNPR<br>(ref: Tigray) | . | . | . | . | -0.103***<br>(0.0204) | -0.0591***<br>(0.0117) | 0.0258***<br>(0.00921) | 0.137***<br>(0.0259) |
| Region – Addis Ababa | . | . | . | . | -0.00114<br>(0.0165) | -0.000649<br>(0.00944) | 0.000283<br>(0.00412) | 0.00150<br>(0.0218) |
| Region – Amhara | . | . | . | . | -0.0616***<br>(0.0227) | -0.0352***<br>(0.0130) | 0.0154**<br>(0.00712) | 0.0814***<br>(0.0298) |
| Region – Oromia | . | . | . | . | 0.0160<br>(0.0406) | 0.00911<br>(0.0232) | -0.00398<br>(0.0102) | -0.0211<br>(0.0537) |
| *Observations* | *1,662* | *1,662* | *1,662* | *1,662* | *1,646* | *1,646* | *1,646* | *1,646* |

**Notes:** \*\*\* p<0.01, \*\* p<0.05, \* p<0.1. Standard errors are in parentheses.

## 5. Discussion and concluding remarks

This paper was motivated by the policy plans of the Ethiopian government to create health insurance schemes to promote universal access to health care. Specifically, in 2011, the government introduced a voluntary CBHI scheme for the rural and informal sectors of urban areas which has since been scaled-up nationwide and proposed a mandatory SHI scheme for the formal sector. Although these schemes target different population segments, in the long run, the intention is to combine the schemes, and establish a large and strong risk pool.

Combining the two schemes requires solidarity across the groups targeted by each of the schemes. Specifically, since it is likely that formal sector employees will have to bear the costs of subsidizing the CBHI scheme (as the premiums will differ while the benefits will be the same), a pertinent policy issue is the willingness of formal sector workers to share health risks and resources with those in the informal sector. Motivated by this policy concern, this study assessed the extent to which formal sector employees were willing to support the CBHI.

Key strengths of the paper include the use of representative data collected by surveying formal sector workers residing in four of the country’s main cities, coverage of both private and public sector employees and a survey experiment to assess the willingness of formal sector workers to support the CBHI. The survey experiment elicited support for the CBHI scheme based on five different cases to which respondents were randomly allocated. These cases differed in terms of the information provided regarding the source of the CBHI subsidy and the benefits associated with the CBHI.

Analysis of the responses showed that, regardless of the information provided on the source of the subsidy or the benefits associated with the CBHI, there was substantial support (76%) for the CBHI. This support cut-across all groups of formal sector workers. While support is high, there is a caveat, as 13% of those surveyed did not respond to the questions on support for the CBHI. Assuming that non-respondents are opposed, cuts down the level of support from 76% to 66%, a lower but still high level of support.

Focusing on those who responded, across the five cases which differed in terms of the information provided regarding the source of the CBHI subsidy and the benefits associated with the CBHI, support for the CBHI ranged from 72.8% in the baseline case to 79.1% (Case five). Although not always estimated precisely, the logit models provide evidence that communicating and disseminating research-based information on the benefits of the CBHI increases the level of support by about 6 percentage points. The responsiveness of respondents to research findings is a useful result as it suggests that it is possible to influence opinions and thereby public policy and generate support for social protection programs such as the CBHI which require cross-group solidarity.

Although the level support is heartening, it is ironic that this high level of support for the CBHI is not matched by support for the introduction of the SHI (Bayked et al., 2023; Zarepour et al., 2023). The reluctance to support the SHI stems from concerns about the costs and skepticism about the benefits, specifically the quality of care available from public health facilities, and the exclusion of private sector health facilities from the ambit of the scheme. In contrast, the support for the CBHI, may well be motivated by the nation-wide scaling-up and sustainability of the CBHI which inspires confidence and signals that the CBHI is functioning satisfactorily.

The estimates showed that knowledge and awareness of the principles of health insurance has a large bearing on the level of support for the CBHI. This is policy-relevant information. Just as in the case of communicating research information, enhancing awareness of the principles of health insurance is likely to enhance support for risk-pooling and cross-subsidies (Shrestha et al., 2020; Sendekie et al., 2024).

While knowledge of the functioning of health insurance enhances support, the lower support for CBHI amongst those who already have health insurance, and are likely to be well-aware of the principles of health insurance, displays that solidarity is clearly not universal. Formal sector workers with health insurance may fear that the continued expansion of the CBHI will increase their financial burden as they must pay for their existing (private) health insurance and may still have to contribute to publicly organized insurance schemes such as the CBHI/SHI.

There are sharp regional variations in the level of support for the CBHI. The lowest levels of support are observed in Tigray region (63%) followed by Oromia (69%) while the three other regions show levels of support that are higher or at the average level of support observed in the sample (75% in Addis Ababa, 79% in Amhara, 88% in SNNPR). The strong support for the CBHI in SNNPR may be attributed to the more agrarian nature of the region and the high levels of satisfaction indicated by CBHI users residing in this region. ^14^ A recent meta-analysis of household-satisfaction with the CBHI showed that at 66.8% SNNPR ranked first while at 49.6% households in in Addis Ababa were most dissatisfied (Worede et al., 2023). However, this metric is unlikely to explain the lower level of support for the CBHI in Oromia (64.1% satisfaction) or Tigray (no evidence). It is also not possible to attribute these variations to differences in awareness of CBHI (no statistically significant difference across regions) or of levels of knowledge of the principles of health insurance (82% of formal sector workers in Tigray had high knowledge of health insurance versus the average of 48.9% in the sample). In the case of the Oromia region, it is most likely that the lower levels of support amongst formal sector workers for such federally supported schemes is motivated by long-standing grievances and lack of trust in the federal government (Kelecha, 2021; Salemot and Metshanda, 2023). The low level of support in the Tigray region is harder to explain as the internal conflict between the Ethiopian government and the Tigray region is more recent and dates from 2018 onwards although, the strong grip of this region on power in the federal government has been diluted since 2012. In any case, while relative support for the CBHI is low in Tigray and Oromia, at 63% and 69% it is not low in absolute terms.

Despite variations across regions which reflects a somewhat fractured polity, there is strong willingness and solidarity among tax-paying employees in major Ethiopian cities to support the continued development of a health insurance scheme for the informal sector. This strong support is not contingent on the information provided in the different cases posed to respondents. The results of this study suggest that, as envisaged by the Ethiopian government (HSFR, 2008; Hanson et al., 2023), in the long run, there is a possibility to create a larger social protection scheme by combining the CBHI and the SHI without substantial friction. While this is promising, it is perhaps cold comfort as the larger health policy challenge remains the resistance to the introduction of the SHI scheme amongst formal sector workers (Zarepour et al., 2023). Unless the concerns underlying the introduction of the SHI are addressed, its introduction and the creation of a national level risk-pooling will remain elusive.

## Data Availability

All data produced in the present study are available upon reasonable request to the authors

## Author contributions

**AM** Survey design, Sampling approach, Data collection, Data curation, Data analysis, Writing – Original draft; **DS** Survey design, Sampling approach, Data collection, Data curation, Writing – Reviewing and editing; **GA** Conceptualization, Survey design, Sampling approach, Data collection, Project administration, Writing – Reviewing and editing; **ZS** Conceptualization, Funding acquisition, Writing – Review and Editing; **AB** Conceptualization, Survey design, Sampling approach, Funding acquisition, Writing – Original Draft

All authors confirm full access to all data used in the study and accept responsibility for the journal submission.

## Data accessibility statement

The data set used for this study is available from the corresponding author.

## Ethics and consent

Informed consent was obtained from respondents prior to data collection. Ethics approval (IDPR/LT-0005/2016) was provided by the Research Ethics Committee of the Institute of Development Policy and Research, Addis Ababa University.

## Appendix Tables

**Table A1.** Potential SHI members based on a 2013 labor force survey and estimates from Ethiopian Health Insurance Agency.

| Sector | Number of employees<br>(Share of total in %) | Sample distribution-<br>*Planned*<br>(Share of total in %) | Sample distribution-<br>*Achieved*<br>(Share in total in %) |
| --- | --- | --- | --- |
| Government worker/civil servant <sup>a</sup> | 1,410,572<br>(32.8) | 690<br>(32.8) | 637<br>(33.2) |
| Public sector enterprise <sup>b</sup> | 424,039<br>(9.9) | 208<br>(9.9) | 191<br>(9.9) |
| Private sector enterprise <sup>b</sup> | 1,789,963<br>(41.7) | 872<br>(41.5) | 757<br>(39.4) |
| NGO workers <sup>b</sup> | 84,808<br>(1.98) | 42<br>(2.0) | 39<br>(2.0) |
| Civil servant/public sector enterprise pensioners <sup>a</sup> | 590,024<br>(13.75) | 289<br>(13.76) | 295<br>(15.4) |
| Total | 4,290,406 | 2,101 | 1,919 |

**Notes:** <sup>a</sup> From EHIA, actual number of employees in May 2016; <sup>b</sup> Estimated from Central Statistical Office, labor force survey

**Table A2.** CBHI scenarios presented to the survey instruments.

|  |
| --- |
| **Case 1** |
| a. Have you heard of the Community Based Health Insurance Scheme which offers health insurance to rural households and those working in the informal sector? (1 = Yes, 2 = No). |
| b. Do you oppose or support this scheme? (1 = strongly opposed, 2 = opposed, 3 = neither oppose nor support, 4 = support, 5 = strongly support). |
| **Case 2** |
| a. Have you heard of the Community Based Health Insurance Scheme which offers health insurance to rural households and those working in the informal sector? (1 = Yes, 2 = No). |
| b. The scheme allows all those who enroll to access health services for a premium of 15 Birr per month. This is the premium that needs to be paid for the entire household. In addition, the government pays a subsidy of 5 Birr per month for each household that enrolls and 10% of indigent households get a complete fee waiver. The costs of this subsidy will be borne by those working in the formal sector/taxpayers. |
| c. Do you oppose or support this scheme? (1 = strongly opposed, 2 = opposed, 3 = neither oppose nor support, 4 = support, 5 = strongly support). |
| **Case 3** |
| a. Have you heard of the Community Based Health Insurance Scheme which offers health insurance to rural households and those working in the informal sector? (1 = Yes, 2 = No). |
| b. The scheme allows all those who enroll to access health services for a premium of 15 Birr per month. This is the premium that needs to be paid for the entire household. In addition, the government pays a subsidy of 5 Birr per month for each household that enrolls and 10% of indigent households get a complete fee waiver. The costs of this subsidy will be borne by international donors. |
| c. Do you oppose or support this scheme? (1 = strongly opposed, 2 = opposed, 3 = neither oppose nor support, 4 = support, 5 = strongly support). |
| **Case 4** |
| a. Have you heard of the Community Based Health Insurance Scheme which offers health insurance to rural households and those working in the informal sector? (1 = Yes, 2 = No). |
| b. The scheme allows all those who enroll to access health services for a premium of 15 Birr per month. This is the premium that needs to be paid for the entire household. In addition, the government pays a subsidy of 5 Birr per month for each household and 10% of indigent households get a complete fee waiver. The costs of this subsidy will be borne by those working in the formal sector/taxpayers. |
| Research finds that almost 50% of those who were offered the insurance have joined the scheme. The use of health care has increased by 50-60 percent and per health care visit, households pay 60% less than they were paying before they had insurance. |
| c. Do you oppose or support this scheme? (1 = strongly opposed, 2 = opposed, 3 = neither oppose nor support, 4 = support, 5 = strongly support). |
| **Case 5** |
| a. Have you heard of the Community Based Health Insurance Scheme which offers health insurance to rural households and those working in the informal sector? (1 = Yes, 2 = No). |
| b. The effectiveness of the scheme has been studied and research finds that almost 50% of those who were offered the insurance have joined the scheme. The use of health care has increased by 50-60 percent and per health care visit, households pay 60% less than they were paying before they had insurance. |
| c. Do you oppose or support this scheme? (1 = strongly opposed, 2 = opposed, 3 = neither oppose nor support, 4 = support, 5 = strongly support). |
| **A follow up question to all cases** |
| For all those who support or strongly support the scheme (that is, response 4 or 5) ask if they are willing to endorse continued government support of the scheme by signing a slip of paper and placing their signature in a box (1 = Yes, 2 = No). |

**Table A3.** Comparing profiles of study participants across cases (Kruskal–Wallis test)

|  | Case 1 | Case 2 | Case 3 | Case 4 | Case 5 | Chi2 | P-value |
| --- | --- | --- | --- | --- | --- | --- | --- |
| Sex of the respondent-male | 0.635 | 0.651 | 0.644 | 0.646 | 0.629 | 0.582 | 0.9651 |
| Age group-less than 30 | 0.272 | 0.299 | 0.265 | 0.286 | 0.273 | 3.339 | 0.5028 |
| Age group-30 - 39 | 0.319 | 0.326 | 0.329 | 0.265 | 0.288 |  |  |
| Age group- 40 - 49 | 0.187 | 0.177 | 0.197 | 0.215 | 0.236 |  |  |
| Age group- 50+ | 0.223 | 0.198 | 0.209 | 0.234 | 0.202 |  |  |
| Education level-No education | 0.013 | 0.012 | 0.009 | 0.012 | 0.004 | 4.654 | 0.3247 |
| Education level-Primary | 0.104 | 0.087 | 0.124 | 0.089 | 0.075 |  |  |
| Education level-Secondary | 0.117 | 0.157 | 0.191 | 0.129 | 0.169 |  |  |
| Education level-Post Secondary | 0.764 | 0.738 | 0.674 | 0.766 | 0.745 |  |  |
| Education level-Other & informal | 0.003 | 0.006 | 0.003 | 0.003 | 0.007 |  |  |
| Marital status-Married | 0.592 | 0.616 | 0.582 | 0.657 | 0.630 | 5.272 | 0.2605 |
| Marital status-Single | 0.281 | 0.270 | 0.282 | 0.259 | 0.249 |  |  |
| Marital status-Divorced | 0.055 | 0.052 | 0.071 | 0.052 | 0.042 |  |  |
| Marital status-Widowed | 0.073 | 0.061 | 0.065 | 0.031 | 0.079 |  |  |
| Religion-Orthodox | 0.813 | 0.799 | 0.775 | 0.783 | 0.789 | 3.344 | 0.5021 |
| Religion-Protestant | 0.148 | 0.142 | 0.166 | 0.173 | 0.170 |  |  |
| Religion-Muslim | 0.031 | 0.052 | 0.047 | 0.037 | 0.030 |  |  |
| Religion -Other religion | 0.008 | 0.006 | 0.012 | 0.006 | 0.011 |  |  |
| Ethnicity-Amhara | 0.462 | 0.453 | 0.448 | 0.450 | 0.519 | 3.988 | 0.4076 |
| Ethnicity-Oromo | 0.178 | 0.159 | 0.211 | 0.168 | 0.199 |  |  |
| Ethnicity-Tigrie | 0.207 | 0.247 | 0.202 | 0.217 | 0.128 |  |  |
| Ethnicity-Guragie | 0.050 | 0.056 | 0.050 | 0.056 | 0.053 |  |  |
| Ethnicity-Other ethnic | 0.102 | 0.085 | 0.089 | 0.109 | 0.102 |  |  |
| Monthly income <sup>a</sup> | 5775.1<br>(4780.9) | 5524.0<br>(4921.1) | 6087.7<br>(9175.0) | 5692.2<br>(7035.5) | 5675.9<br>(6210.2) | 0.340 | 0.8510 |
| Own house | 0.383 | 0.372 | 0.363 | 0.425 | 0.386 | 1.908 | 0.7526 |
| Household size - 1 | 0.254 | 0.250 | 0.241 | 0.255 | 0.255 | 0.814 | 0.9366 |
| Household size – 2-3 | 0.334 | 0.314 | 0.318 | 0.305 | 0.326 |  |  |
| Household size – 4-5 | 0.306 | 0.375 | 0.329 | 0.348 | 0.300 |  |  |
| Household size – 6+ | 0.106 | 0.061 | 0.112 | 0.092 | 0.120 |  |  |
| Any illness in the household | 0.280 | 0.294 | 0.341 | 0.246 | 0.285 | 6.917 | 0.1403 |
| Household outpatient care use | 0.264 | 0.267 | 0.324 | 0.268 | 0.255 | 4.427 | 0.3513 |
| Household member smokes | 0.037 | 0.021 | 0.035 | 0.028 | 0.030 | 1.671 | 0.7961 |
| Household member drinks | 0.501 | 0.472 | 0.478 | 0.461 | 0.445 | 1.915 | 0.7514 |
| Household member has *khat* | 0.031 | 0.053 | 0.041 | 0.037 | 0.042 | 1.808 | 0.7711 |
| Insurance membership | 0.522 | 0.496 | 0.500 | 0.500 | 0.496 | 0.686 | 0.9530 |
| Sector-Public sector | 0.339 | 0.358 | 0.356 | 0.345 | 0.363 | 0.575 | 0.9658 |
| Sector-Public enterprises | 0.096 | 0.102 | 0.100 | 0.120 | 0.101 |  |  |
| Sector-Private and NGO | 0.438 | 0.410 | 0.409 | 0.409 | 0.419 |  |  |
| Sector-Pension | 0.127 | 0.131 | 0.135 | 0.126 | 0.116 |  |  |
| Region – Tigray | 0.174 | 0.195 | 0.153 | 0.169 | 0.079 | 5.059 | 0.2813 |
| Region – Addis Ababa | 0.518 | 0.480 | 0.509 | 0.486 | 0.513 |  |  |
| Region – Amhara | 0.124 | 0.134 | 0.138 | 0.132 | 0.172 |  |  |
| Region – Oromia | 0.021 | 0.026 | 0.024 | 0.022 | 0.019 |  |  |
| Region – SNNPR | 0.163 | 0.166 | 0.176 | 0.191 | 0.217 |  |  |
| N | 386 | 344 | 340 | 325 | 267 |  |  |

**Notes:** <sup>a</sup> Standard deviations are in parentheses. Differences in mean income by group is based on an analysis of variance (ANOVA) test. Of 1919 respondents, 257 did not respond to the question on supporting CBHI.

**Table A4.**
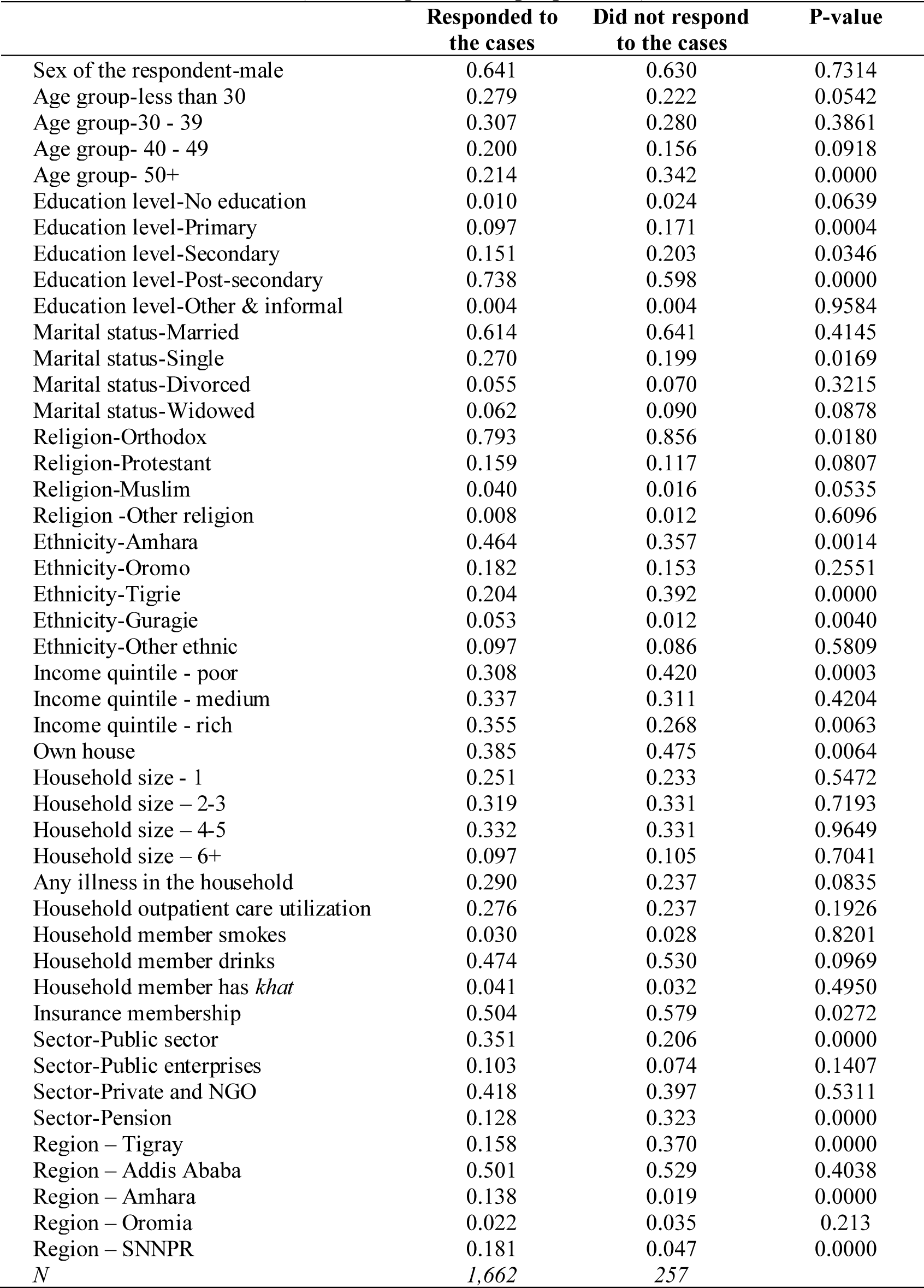
Comparison of who responded to the CBHI case questions and those who did not (Two-sample test of proportions)

## Footnotes

1 The SHI scheme targets public and private formal sector workers, NGOs employees, as well as pensioners. While the two schemes, CBHI and SHI, target different groups, the long-term plan is to combine the two programs to create a larger risk pool at the national level (HSFR, 2008).

2 Ethiopian Health Insurance Service (EHIS) 2023/24, annual performance report.

3 Exceptions are health insurance schemes in Ghana and Rwanda.

4 A design team reviewed practices around the world and study tours were undertaken to understand the schemes operating in Ghana, Mexico, Rwanda, Senegal, China, Vietnam, and Thailand (HSFR, 2009; Abt Associates, 2015). Additionally, feasibility studies were conducted in four regional states - Amhara, Oromia, SNNP and Tigray.

5 The revision of the scheme focused mainly on measures to enhance financial sustainability by adjusting membership contributions and expanding coverage of the program (Segahu, 2018).

6 At scheme inception, the decision to consider enrolling in the CBHI was made at the village/community level based on a simple majority. Once a community decided that the CBHI would be on offer, all households were expected to join the scheme. In practice, there was no enforcement mechanism and households joined the scheme on a voluntary basis.

7 Household level enrolment is intended to reduce adverse selection. Furthermore, after registering, new members need to wait for one month before they can use services. At inception, as an additional measure to diversify risk and create larger risk pools the scheme was only launched in a district if (intended) enrolment reached 30% of eligible fee-paying households. This threshold was revised to 50% in July 2016 (Abt Associates, 2015; Abdilwohab et al., 2021) and to 80% as of January 2024 (ANRS, 2024).

8 Since 2016, in all regions, the premium is expected to be paid in one go on an annual basis. However, in practice this varies across districts.

9 Based on a household panel survey, CSS and World Bank (2023), report that the average annual household consumption expenditure per adult was 25,000 Birr in urban areas and 14,000 Birr in rural areas while average household size was 4.2 in urban areas and 5.2 in rural area. Based on these data, in urban areas the CBHI premium lies between 0.85-3.3% of the average urban household expenditure and in rural areas the corresponding figure is 1.09-1.15%. CSS and World Bank. (2023). Ethiopia Socioeconomic Panel Survey (ESPS) Report - Wave 5, 2021/22.Ethiopian Statistical Service and World Bank: Addis Ababa http://www.statsethiopia.gov.et/wp-content/uploads/2024/01/2021-22-Survey-Report.pdf

10 The bulk (70%) of the fee-waiver subsidy is financed by regional governments and the remainder is sourced from district governments.

11 Regional and city level risk pools have been established in Harari regional state and Dire Dawa city and since 2018, zonal level risk pools have been implemented in Amhara, Oromia, Sidama, and Central Ethiopia (HSFR, 2018; EHIA, 2020; USAID, 2021). Ethiopia has a federal system of government with various administrative structure. It is divided into 11 regional states and 2 chartered cities (Addis Ababa and Dire Dawa). Regions are further subdivided into zones and woredas (districts).

12 For outpatient services, a majority of formal sector employees (56%) opt for care from private health facilities while for inpatient services the corresponding figure is 37.5% (Zarepour et al., 2023).

13 As mentioned in the previous section, the outcome variable was measured on a 5-point Likert scale. However, the proportion of individuals in the categories strongly opposed and opposed were relatively small and hence we combined the two responses.

14 The country’s SNNP region is predominantly agrarian and is characterized by strong social networks/social capital (Endris et al., 2017; Noguchi, 2018) which may have contributed to the strong support for CBHI amongst respondents from SNNPR.

